# Effect of Electroacupuncture on Delayed-Onset Muscle Soreness Induced by Eccentric Exercise in Young Untrained Men: Comparison of Contralateral and Ipsilateral Stimulation

**DOI:** 10.1101/2024.04.16.24305776

**Authors:** Shoichi Komine, Ikuru Miura, Teng Hu, Akihiro Ogata, Shinsuke Tamai, Katsuyuki Tokinoya, Takehito Sugasawa, Sechang Oh, Shota Wada, Kazuhiro Takekoshi, Go Ito, Tomonori Isobe, Takashi Matsui, Hajime Ohmori

**Affiliations:** Department of Acupuncture and Moxibustion, Faculty of Health Care, Teikyo Heisei University, Toshima-ku, Tokyo, Japan; Research Institute of Oriental Medicine, Teikyo Heisei University, Toshima-ku, Tokyo, Japan; Institute of Medicine, University of Tsukuba, Tsukuba, Ibaraki, Japan; Faculty of Sports and Health Science, Fukuoka University, Fukuoka, Japan; Institute for Physical Activity, Fukuoka University, Fukuoka, Japan; Graduate School of Human Health Sciences, Tokyo Metropolitan University, Hachioji, Tokyo, Japan; Institute of Human Sciences, University of Tsukuba, Tsukuba, Ibaraki, Japan; Department of Sport Science and Research, Japan Institute of Sports Sciences, Kita-ku, Tokyo, Japan; Embodied Wisdom Division, Center for Liberal Education and Learning, Sophia University, Chiyoda-ku, Tokyo, Japan; Faculty of Rehabilitation, R Professional University of Rehabilitation, Tsuchiura, Japan; Graduate School of Comprehensive Human Sciences, University of Tsukuba, Tsukuba, Ibaraki, Japan; Oriental Medicine Research Center, Kitasato University, Minato-ku, Tokyo, Japan; Institute of Health and Sport Sciences, University of Tsukuba, Tsukuba, Ibaraki, Japan; Department of Sports and Health Management, Faculty of Business Information Sciences, Jobu University, Isesaki, Gunma, Japan; University of Tsukuba, Tsukuba, Ibaraki, Japan

## Abstract

**Background:** Electroacupuncture stimulation is sometimes used to prevent and treat delayed onset muscle soreness (DOMS) induced by exercise; however, the effect is unclear. Using a human biceps brachii muscle pain induction model, we investigated the effect of contralateral electroacupuncture (CEA) versus ipsilateral electroacupuncture (IEA) stimulation on DOMS and muscle injury markers.

**Methods:** Nineteen young men were randomly assigned to receive either CEA or IEA. All participants performed eccentric exercise using the biceps brachii muscle of the nondominant arm to induce muscle damage. Electroacupuncture stimulation (1.5 Hz, 15 minutes) was applied to the dominant arm in the CEA group and the nondominant arm in the IEA group. Electroacupuncture stimulation was applied in both groups from 7 days before exercise to 4 days after exercise. All variables were analyzed at the following seven timepoints: baseline, immediately before exercise, immediately after exercise, and on days 1–4 after exercise.

**Results:** Palpation pain was significantly lower in the IEA group than the CEA group at 72 and 96 hours after exercise. The decrease in the elbow range of motion after exercise was significantly suppressed by electroacupuncture stimulation. The muscle injury markers (creatine kinase, lactate dehydrogenase) increased following exercise, but these changes were not significantly influenced by IEA stimulation. IEA suppressed exercise-induced oxidative stress at 72 hours after exercise.

**Conclusion:** This study suggests that the application of IEA before and after eccentric exercise effectively reduces DOMS. Electroacupuncture might suppress increases in oxidative stress elicited by eccentric exercise.

## 1 Introduction

Exercise is beneficial for overall health and is known to improve muscle strength, weight loss, mental health, psychological and cognitive function, sleep disorders, type 2 diabetes, and cardiac diseases^1-5^. However, when performed by people who are unfamiliar with high-intensity exercise, exercise can induce muscle soreness and damage. Muscle soreness that occurs several days after exercise is called delayed onset muscle soreness (DOMS) and can be observed as pain during exercise or skeletal muscle palpation^6^. DOMS can lead to a discrepancy between the perceived effort of muscle strength exertion and the actual muscle strength^7^. Additionally, DOMS induces muscle weakness and altered proprioception, increasing the risk of secondary impairments such as decreased exercise performance and the potential for falls^7-9^. Therefore, the inhibition of DOMS is crucial.

DOMS and muscle damage are subjectively assessed using visual scales, and objectively evaluated by measuring changes in the blood concentrations of enzymes such as creatine kinase and lactate dehydrogenase^10^. In addition to conventional markers of muscle damage, blood cell-free DNA (cfDNA) collected immediately after high-intensity exercise has recently been used to measure exercise-induced changes and as a new biomarker of muscle damage^11^. Furthermore, the post-exercise-specific cfDNA (spcfDNA) efflux has been highlighted as a potentially useful marker of muscle damage^12^.

Acupuncture is a traditional Eastern therapeutic method that is often used to treat skeletal muscle soreness. Several studies have examined the effects of acupuncture stimulation on DOMS. Itoh et al. reported that DOMS is suppressed when acupuncture is applied to areas with tenderness induced by eccentric exercise^13^, and a recent meta-analysis concluded that general acupuncture is effective in alleviating DOMS^14^. However, the effectiveness of electroacupuncture for DOMS remains unclear. Furthermore, there are cases in which it may not be possible to apply acupuncture on the injured side. Such cases may require opposing acupuncture, also known as healthy-side acupuncture or contralateral acupuncture, which is one of the nine acupuncture methods recorded in the classic Chinese medicine textbook Huangdi Neijing (The Yellow Emperor’s Canon of Internal Medicine). Contralateral acupuncture is applied to the opposite side to the painful area^15-18^. However, the literature on direct comparisons of the effect of different acupuncture methods on DOMS is limited.

To address this knowledge gap, we examined the effects of direct ipsilateral electroacupuncture (IEA) versus contralateral electroacupuncture (CEA) on the occurrence of DOMS after eccentric exercise. We also measured conventional muscle damage markers, oxidative stress markers, and spcfDNA. We hypothesized that the application of IEA before and after eccentric exercise would decrease the incidence of DOMS and reduce oxidative stress after eccentric exercise.

## 2 Materials and Methods

### 2.1 Ethics Statement

The study protocol was approved by the ethics committees of the University of Tsukuba, Japan (approval number: Tai019-75) and Teikyo Heisei University (approval number: 2020-100) and followed the ethics principles of the 7th revision (2013) of the Declaration of Helsinki. Participants provided written informed consent and authorization for the disclosure of protected health information before enrolling in the study.

### 2.2 Experimental Design

The study cohort comprised 19 healthy men with no knowledge of or experience with acupuncture and no exercise habits, who had not smoked or used medication in the preceding 3 months. Participants were instructed to refrain from (i) massaging their upper arm, (ii) strenuous exercise, (iii) alcohol consumption, (iv) taking medication, (v) taking supplements, and (vi) receiving acupuncture or electroacupuncture treatment during the experiment. Numbers were randomly generated by a computer and aligned with each participant for randomization to divide the participants into the CEA group and the IEA group. The experimental schedule is shown in Figure 1A. Before the experiment, the height, body weight, body mass index, percentage fat, and muscle mass were measured using a 4-Compartment Model Method body composition analyzer (RD-803L, Tanita, Tokyo, Japan). To ensure that the degree of muscle soreness after exercise was approximately equivalent between individuals, the maximal voluntary contraction (MVC) of the biceps of the nondominant arm was measured using the Biodex System 4 (Sakai Med. Co., Tokyo, Japan). Briefly, the maximum torque was analyzed while the participants performed isometric contractions in the sitting position with the shoulder and elbow joints both at 90 degrees of flexion. The MVC was measured twice, and the average MVC value was recorded. The average MVC was then corrected according to the length of the participant’s forearm. To exclude the influence of the MVC measurement, a 2-week washout period was applied. After 2 weeks (i.e., at the Pre timepoint), all participants assembled in the laboratory at 8:00 A.M. We measured the muscle soreness and the elbow joint angle and range, and collected blood. Each participant then underwent electroacupuncture of the dominant arm in the CEA group and the nondominant arm in the IEA group. On the exercise day (7 days after Pre), all participants assembled in the laboratory at 8:00 A.M. Both groups underwent biceps brachii evaluation and blood sample collection within 30 minutes before starting eccentric exercise (i.e., at the Ex-Pre timepoint) and immediately after the exercise (i.e., at the Ex-Post timepoint). At 24, 48, and 72 hours after the exercise session, the participants underwent measurements, blood sampling, and electroacupuncture. At 96 hours after the exercise, only measurements were performed, and the participants did not receive electroacupuncture. All measurements were performed by the same evaluator, who was blinded to the group allocation.

**Fig. 1.**
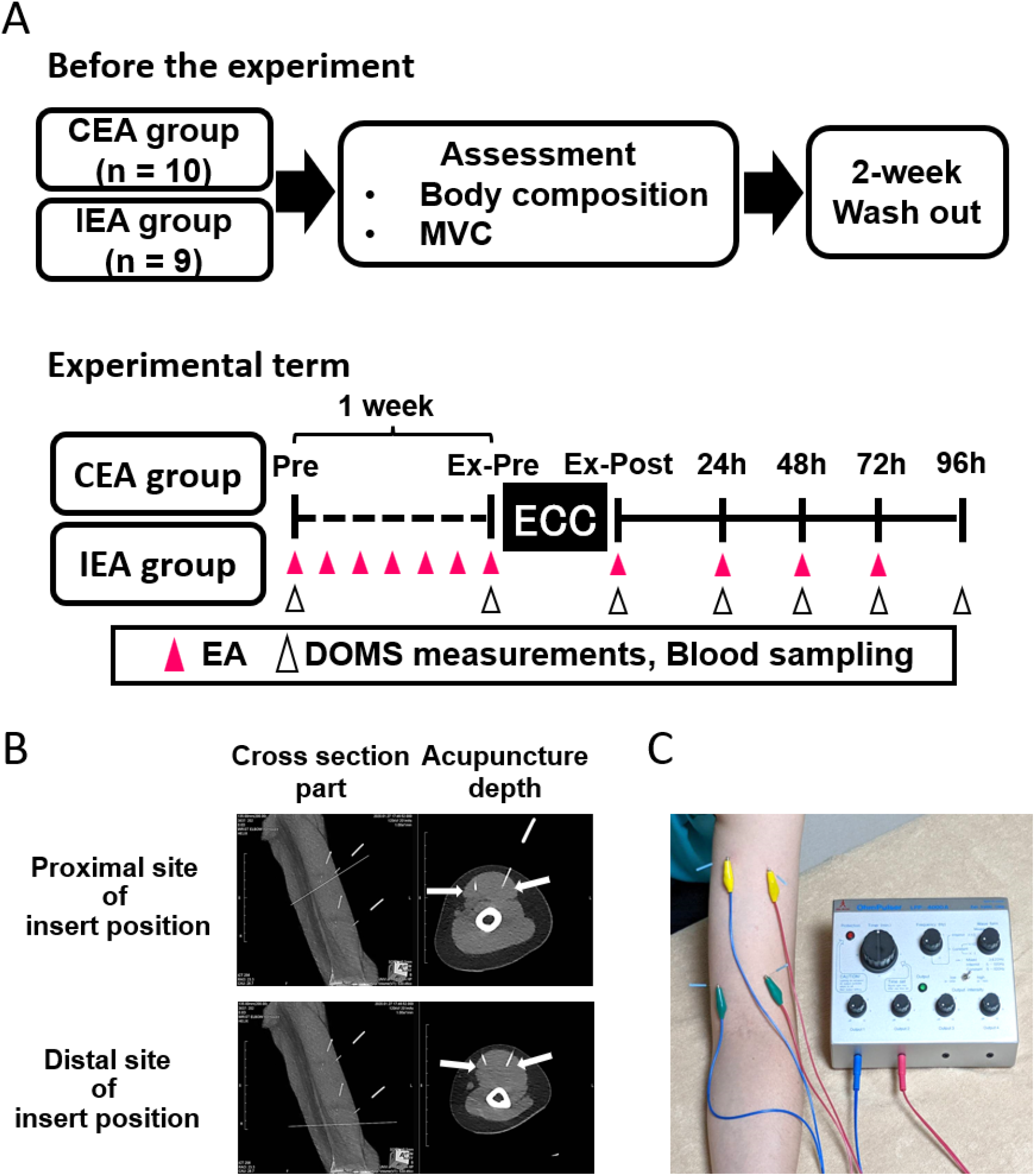
Experimental flow and electroacupuncture stimulation (A) Experimental flow. Pink arrowheads indicate electroacupuncture stimulation. White arrowheads indicate the timepoints at which DOMS evaluation and blood sample collection were performed. Electroacupuncture was performed after the measurement. (B) Electroacupuncture stimulation sites in the distal and proximal biceps brachii (magnetic resonance imaging). White lines indicate acupuncture needles. Arrows indicate the needle tips. (C) Position of the participant during electroacupuncture stimulation. CEA, contralateral electroacupuncture; DOMS, delayed onset muscle soreness; ECC, eccentric exercise; IEA, ipsilateral electroacupuncture; MVC, maximum voluntary contraction.

### 2.3 Dietary Analysis

Dietary assessment of the participants was conducted by qualified personnel using the 24-hour recall method^16, 17^ Subjects were asked to record their dietary intake, including three meals (breakfast, lunch, and dinner), snacks, and beverages, for the first 3 days after exercise. Participants who provided us with photographs of their daily diet were given a 15-min nutritional interview session, during which the information was reviewed. Total amount, amino acids, total fat, total carbohydrate, available carbohydrates, total fiber and energy intake of dietary contents were estimated using the nutrition calculation software included in the Standard Tables of Food Composition in Japan 2019, 7th edition (Ishiyaku Publishers, Inc., Tokyo Japan). The calculations were performed by a nationally certified dietitian.

### 2.4 Electroacupuncture

Electroacupuncture stimulation was performed 6 days before the exercise day, before and after eccentric exercise on the same day, and 3 days after exercise (Fig. 1A). Before the experiment, we explained to the participants that the IEA and CEA techniques were being compared to eliminate the placebo effect. Electroacupuncture was applied to the dominant arm (the arm not experiencing DOMS) in the CEA group and the nondominant arm (the arm experiencing DOMS) in the IEA group. The acupuncture needles (0.2 mm diameter; SEIRIN Co., Ltd., Shizuoka, Japan) were inserted to a depth of approximately 20 mm. Computed tomography images were obtained using a Brilliance iCT scanner (Philips Electronics Ltd., Amsterdam, the Netherlands; Fig. 1B). Four needles were placed at the distal and proximal ends of the long and short heads of the biceps brachii muscle. Electric stimulation was applied to the biceps brachii muscle in the longitudinal direction at a frequency of 1 Hz for 15 minutes using an Ohm Pulser LFP-4000A (Zen-Iryoki Co., Ltd., Fukuoka Japan; Fig. 1C). Every pulse waveform was a 250-µs asymmetric dipole pulse wave. This frequency was determined by referring to stimuli used in previous studies ^19^. Because the sensation of pain varies between individuals, the intensity of electrical stimulation was set to a level at which the participant felt comfortable (approximately 2–4 V), and the degree of contraction was set to a level where minute contractions were observed at regular intervals. The stimulus intensity was adjusted to a level that was confirmed as comfortable by the participant, with the intensity being decreased when pain was felt. Acupuncture stimulation was performed by a Japanese national licensee who was unaware of the group allocations.

### 2.5 Exercise Protocol

The exercise, blood sampling, and muscle assessments were performed on an empty stomach early in the morning (06:00–08:30). Participants performed eccentric exercise of the brachial flexor muscle group of the nondominant arm^10^. On the exercise day (Ex-Pre), we collected blood samples, evaluated the range of motion, and measured muscle soreness. During the exercise, the participants were seated with their lumbar spine flexed at 45 degrees and their chest supported. Starting with the shoulder and elbow joints flexed at 90 degrees, they slowly extended the elbow joint to apply the load. The exercise intensity was set to 70% MVC, and six sets of 5 seconds × 5 repetitions were performed. There was a 2-minute break between sets. The exercise timing relied on an electronic metronome (60 beats/minute), and participants who were unable to maintain the rhythm by themselves were assisted. After each 5-second eccentric contraction, it took 3 seconds to achieve the correct arm position before starting the next 5-second eccentric contraction. All contractions started at the same position while the participant was relaxed.

### 2.6 Muscle Soreness Assessment

Subjective biceps muscle soreness was evaluated using a visual analog scale, which consisted of a 100 mm straight line with “no pain” at the left end and “extreme pain” at the right end. Muscle soreness was assessed before exercise, immediately after exercise, and during the next 4 days. Two types of muscle soreness evaluation were performed: palpation and spontaneous maximal extension^10^. Muscle soreness on palpation was evaluated by palpating the biceps muscle belly for 3 s. The subjects extended their elbow joint to assess muscle soreness during spontaneous maximal extension. The visual analog scale was calculated as the score after deducting the Ex-Pre score. The same researcher evaluated all subjects to reduce assessment variability. The evaluation was performed by a third party who was unaware of the grouping.

### 2.7 Joint angle Evaluation

The elbow joint range, which reflects muscle soreness and damage, was measured using a goniometer (Takase Med, Tokyo, Japan). Starting from the lateral superior condyle of the humerus, we aligned both axes of the goniometer with the acromion and radial styloid process and assessed the resting angle as well as maximum extension and flexion within a pain-free range. Active motion by the subject was used to measure the range of the elbow joint. The elbow joint range was calculated as the difference between the maximum extension and flexion angles. The measured angles were calculated as the angle after deducting the Pre angle. The range was measured twice by a third party who was not aware of the grouping, and the average value was used.

### 2.8 Blood Parameters

Blood samples were collected from the antecubital veins from the opposite arm for electroacupuncture stimulation and assessed by the Tsukuba i-Laboratory LLP for creatine kinase, lactate dehydrogenase, aldolase, and aspartate aminotransferase^17, 19-23^. The serum samples were stored at –80 °C until further analysis.

### 2.9 Oxidative Stress Assessment

Next, we measured the levels of serum malondialdehyde (MDA), a lipid peroxidation product and an indicator of eccentric exercise-induced oxidative stress. Measurements were performed using the 2-thiobarbituric acid reactive substances (TBARS) assay kit (Cayman Chemical, Ann Arbor, MI USA) following the manufacturer’s standard protocol. Briefly, the absorbance at 540 nm of the MDA-thiobarbituric acid adduct, formed by the reaction of MDA and thiobarbituric acid under high temperature (90–100 °C) and acidic conditions, was measured using a Varioskan microplate reader (Thermo Fisher Scientific, Waltham, MA, USA).

### 2.10 Blood spcfDNA measurement

To analyze post-exercise-specific cfDNA (spcfDNA) flow out of the skeletal muscle, Serum cfDNA was extracted using the Plasma/Serum Cell-Free Circulating DNA Purification Mini Kit (Norgen Biotek Inc., Thorold, ON, Canada) following the manufacturer’s instructions. Then, the spcfDNA sequence (spcfDNA-1) identified by Sugasawa et al. and TaqMan qPCR primers and probes were designed using the Primer-BLAST web tool (National Library of Medicine, Bethesda, MD, USA)^12^. The nucleotide sequences were as follows: spcfDNA-1 (forward: TCTTGTGGCCTTCGTTGGAA, reverse: ATTGACCTCAAAGCGGCTGA, probe: 56-FAM/ATTGACCTC/ZEN/AAAGCGGCTGA/3IABkFQ). The DNA extracted from human fibroblasts (JCRB 1006.7, JCRB Cell Bank; original developers: Kouchi and Namba) was used to create a calibration curve for absolute cfDNA quantification (calibration curve R^2^ > 0.99). All measurements were performed in duplicate.

### 2.11 Statistical Analysis

Statistical analysis was performed using IBM SPSS Statistics for Windows, Version 29.0 (IBM Corp., Armonk, NY, USA). All data are expressed as mean ± standard deviation. Within-group comparisons between various sampling points were performed using a repeated measures two-way analysis of variance (ANOVA) followed by Tukey’s correction for multiple comparisons. Between-group comparisons were performed using the Student’s t-test. Statistical significance was set at *P* < 0.05.

## 3 Results

### 3.1 Body Composition and Dietary Intake After Eccentric Exercise

Body composition and maximal strength of the nondominant arm did not differ between the two groups (Table 1). The two groups also had similar total caloric, amino acid protein, fat, and carbohydrate intakes, as estimated from the meals consumed during the experiment (Table 2).

**Table 1.**
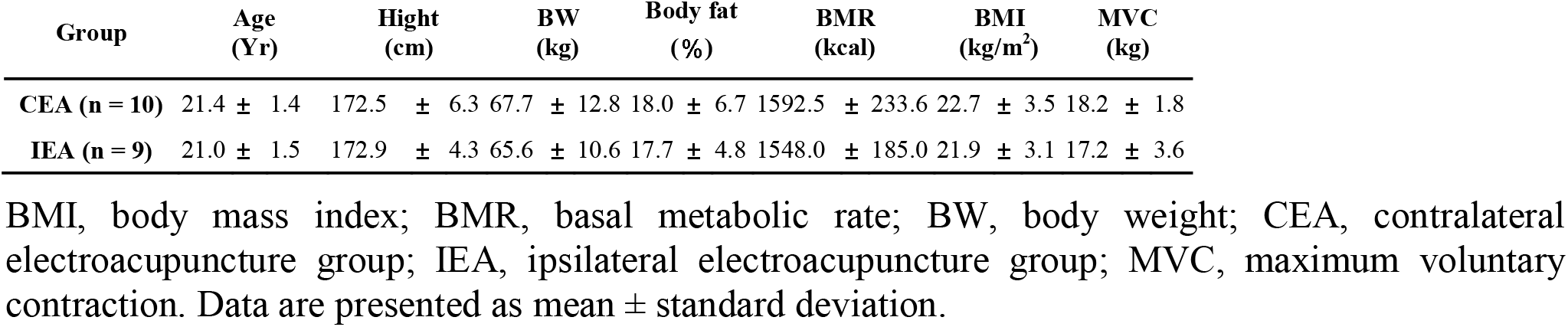
Participant information.

| Group | Age<br>(Yr) | Hight<br>(cm) | BW<br>(kg) | Body fat<br>(%) | BMR<br>(kcal) | BMI<br>(kg/m<sup>2</sup>) | MVC<br>(kg) |
| --- | --- | --- | --- | --- | --- | --- | --- |
| CEA (n = 10) | 21.4 ± 1.4 | 172.5 ± 6.3 | 67.7 ± 12.8 | 18.0 ± 6.7 | 1592.5 ± 233.6 | 22.7 ± 3.5 | 18.2 ± 1.8 |
| IEA (n = 9) | 21.0 ± 1.5 | 172.9 ± 4.3 | 65.6 ± 10.6 | 17.7 ± 4.8 | 1548.0 ± 185.0 | 21.9 ± 3.1 | 17.2 ± 3.6 |

BMI, body mass index; BMR, basal metabolic rate; BW, body weight; CEA, contralateral electroacupuncture group; IEA, ipsilateral electroacupuncture group; MVC, maximum voluntary contraction. Data are presented as mean ± standard deviation.

**Table 2.** Estimated daily dietary content during the experiment.

| Group | Amount<br>(g) | Calorie<br>(kcal) | Protein<br>(g) | Fat<br>(g) | Carbohydrate<br>(g) | Available<br>Carbohydrates<br>(g) | Total Fiber<br>(g) |
| --- | --- | --- | --- | --- | --- | --- | --- |
| CEA (n = 10) | 1684.3 ± 459.9 | 2390.1 ± 531.3 | 90.8 ± 19.3 | 71.7 ± 22.1 | 302.7 ± 63.2 | 268.9 ± 68.8 | 16.1 ± 4.8 |
| IEA (n = 9) | 1416.3 ± 457.8 | 2222.6 ± 463.7 | 76.9 ± 26.6 | 69.4 ± 30.7 | 310.0 ± 53.3 | 284.3 ± 58.1 | 16.8 ± 3.7 |

The 24-hour recall method was used. CEA, contralateral electroacupuncture group; IEA, ipsilateral electroacupuncture group. Data are presented as mean ± standard deviation.

### 3.2 Muscle Soreness Assessment

In the CEA group, pain on palpation was significantly increased at 24 hours after exercise and remained elevated for 96 hours compared with the Pre levels (Fig. 2A); in contrast, the IEA group showed increased pain compared with the Pre levels only at 24 and 48 hours after exercise. Palpation pain at 72 and 96 hours after exercise was significantly lower in the IEA group than the CEA group. We found simple main effects of timepoint and group for pain felt during elbow extension after the exercise session. No interaction was observed (Fig. 2B). Figure 2C and 2D shows heat maps of the visual analog scale values for pain on palpation and extension compared with the Pre values for all participants.

**Fig. 2.**
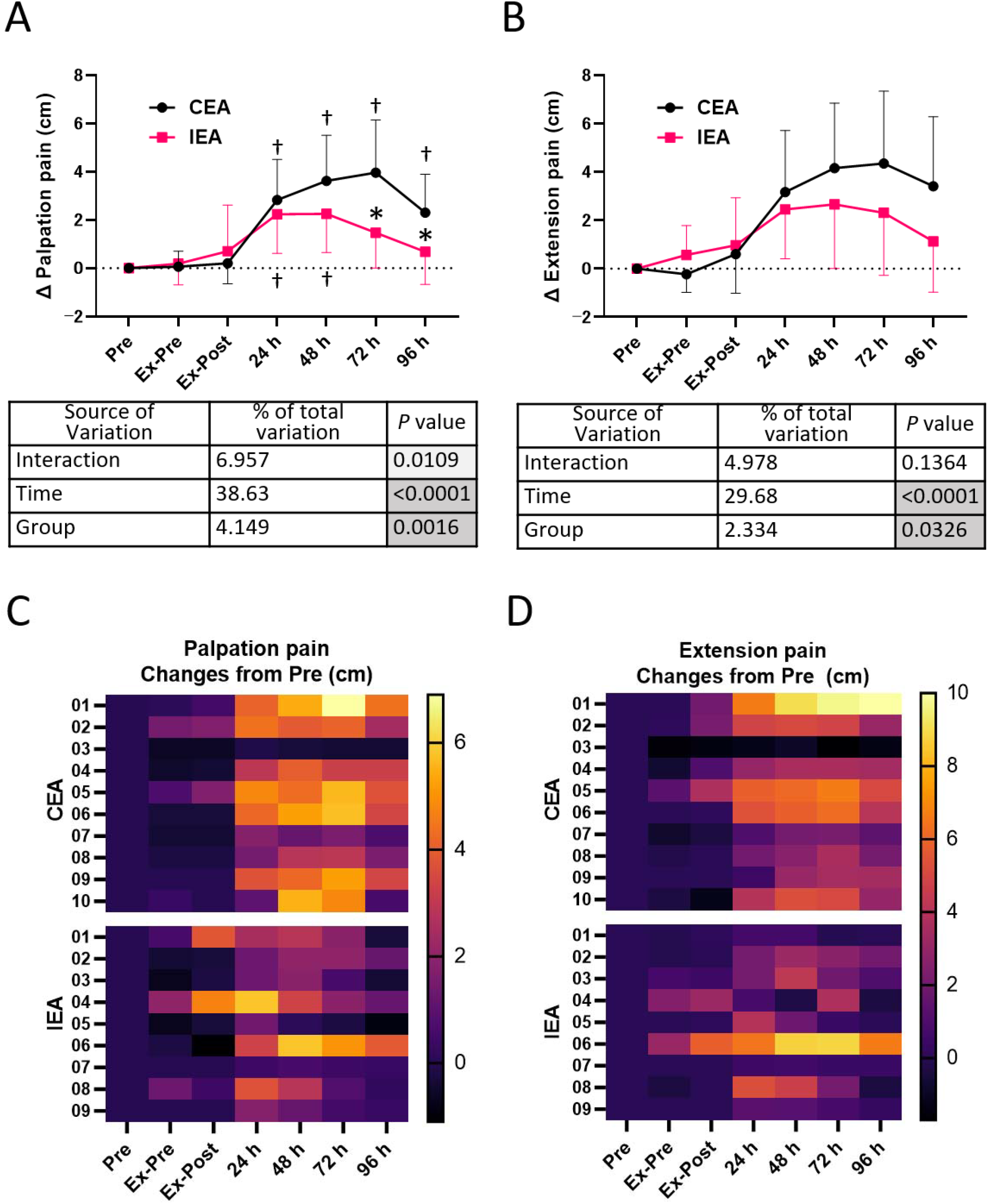
Changes in subjective muscle soreness Evaluated using the visual analog scale. Data are shown as changes from the Pre measurements. Time course of pain on palpation (A) and during extension (B). Heat maps of pain on palpation (C) and during extension (D). Each participant is identified by a number. Asterisks indicate significant differences between the groups (P < 0.05). Daggers indicate significant differences within the group compared with the Pre assessment (P < 0.05). Data are shown as mean ± standard deviation and were statistically compared using a two-way ANOVA followed by Tukey’s post hoc test for multiple group comparisons. In the table, the gray highlighted p-values indicate significant differences. CEA, contralateral electroacupuncture group; IEA, ipsilateral electroacupuncture group; Pre, before the experiment; Ex-Pre, before exercise; Ex-Post, immediately after exercise; 24, 48, 72, and 96 h: 24, 48, 72, and 96 hours after exercise, respectively. n□=□10 in the CEA group, n = 9 in the IEA group.

### 3.3 Elbow Joint Angle Evaluation

To analyze DOMS, we measured the elbow joint angle at which pain was induced. Specifically, we calculated the resting elbow joint angle (Fig. 3A), flexion angle from rest to pain sensation (Fig. 3B), extension angle from rest to pain sensation (Fig. 3C), and painless elbow joint range (Fig. 3D). We found a simple main effect of timepoint and group for pain during elbow extension after the exercise session. No interaction was observed.

**Fig. 3.**
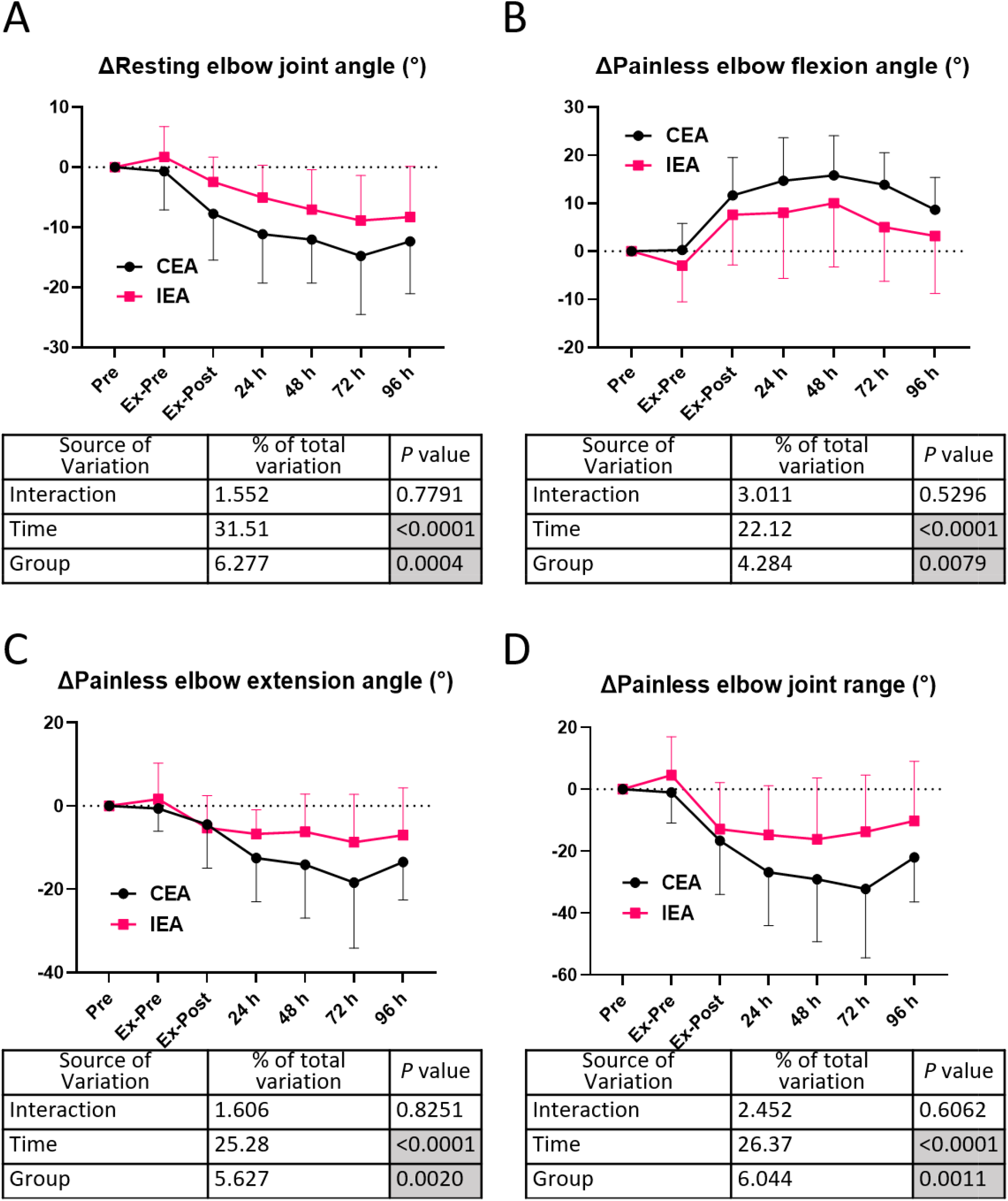
Elbow joint angle and range Resting elbow joint angle and painless elbow joint range. Data are shown as changes from the Pre measurements. Resting elbow joint angle (A). Painless elbow joint flexion (B) and extension (C) angles. Painless elbow joint range (D). Data are presented as mean ± standard deviation and were statistically compared using a two-way ANOVA. CEA, contralateral electroacupuncture group; IEA, ipsilateral electroacupuncture group; Pre, before the experiment; Ex-Pre, before exercise; Ex-Post, immediately after exercise; 24, 48, 72, and 96 h: 24, 48, 72, and 96 hours after exercise, respectively. n□=□10 in the CEA group, n = 9 in the IEA group.

### 3.4 Skeletal Muscle Injury Markers

Changes in the blood concentrations of skeletal muscle damage markers are shown in Figure 4A–D. We found a simple main effect of timepoint for the concentrations of creatine kinase, aspartate aminotransferase, and aldolase after the exercise session. No simple main effect of group was observed, and we found no interaction.

**Fig. 4.**
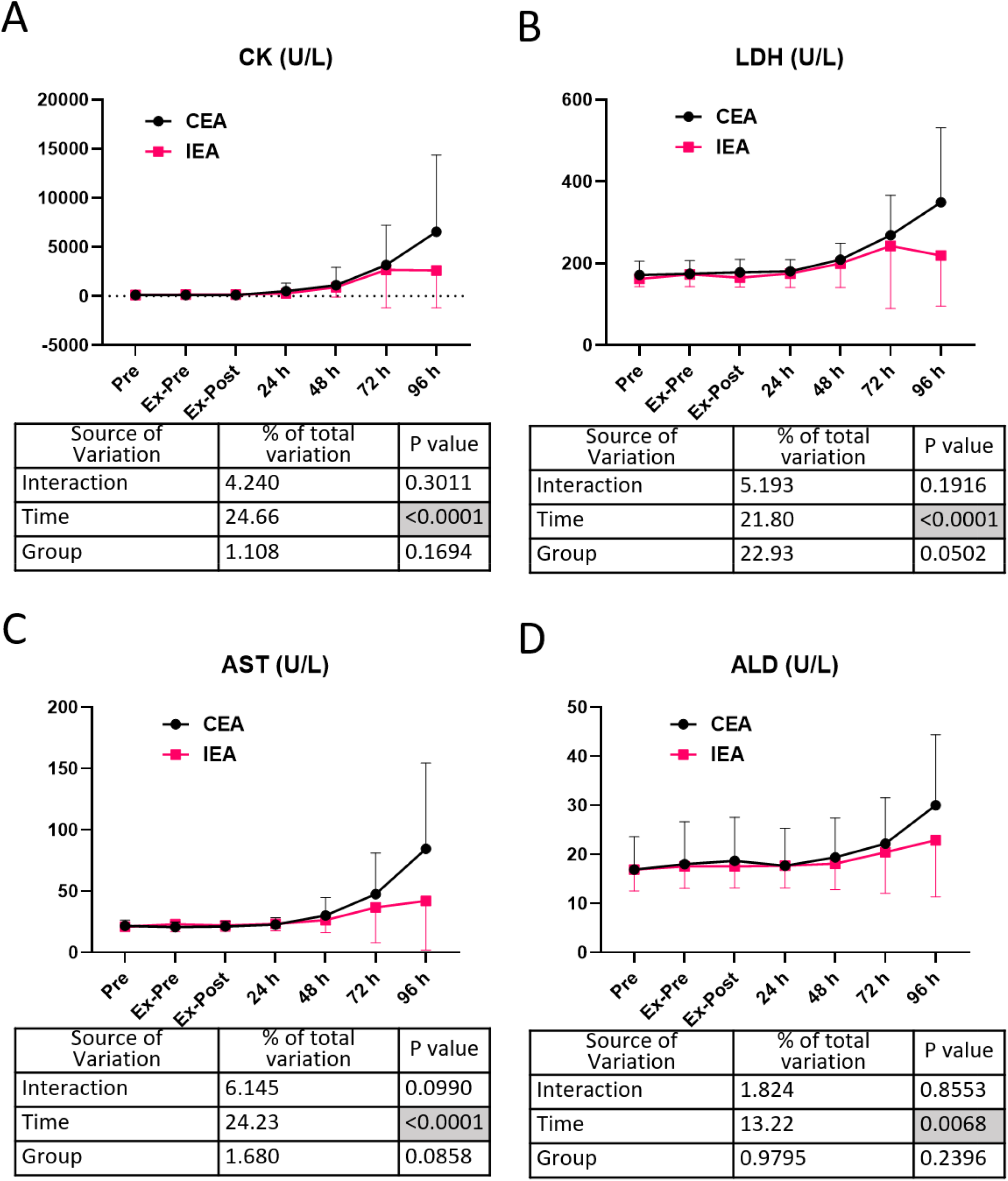
Time course of muscle damage markers Changes in the serum concentrations of muscle injury markers. (A) CK, (B) LDH, (C) AST, and (D) ALD. Data are presented as mean ± standard deviation and were statistically compared using a two-way ANOVA. CEA, contralateral electroacupuncture group; IEA, ipsilateral electroacupuncture group; CK, creatine kinase; LDH, lactate dehydrogenase; AST, aspartate aminotransferase; A D, aldolase; Pre, before experiment; Ex-Pre, before exercise; Ex-Post, immediately after exercise; 24, 48, 72, and 96 h: 24, 48, 72, and 96 hours after exercise, respectively. n□=□10 in the CEA group, n = 9 in the IEA group.

### 3.5 Oxidative stress marker and post-exercise-specific cell-free DNA concentration

To analyze markers involved in DOMS, we measured an oxidative stress marker (MDA) and spcfDNA (Fig. 5). The MDA concentration at 72 hours after exercise was significantly lower in the IEA group than the CEA group (Fig. 5A). The spcfDNA value immediately after exercise was significantly lower in the IEA group than the CEA group (Fig. 5B).

**Fig. 5.**
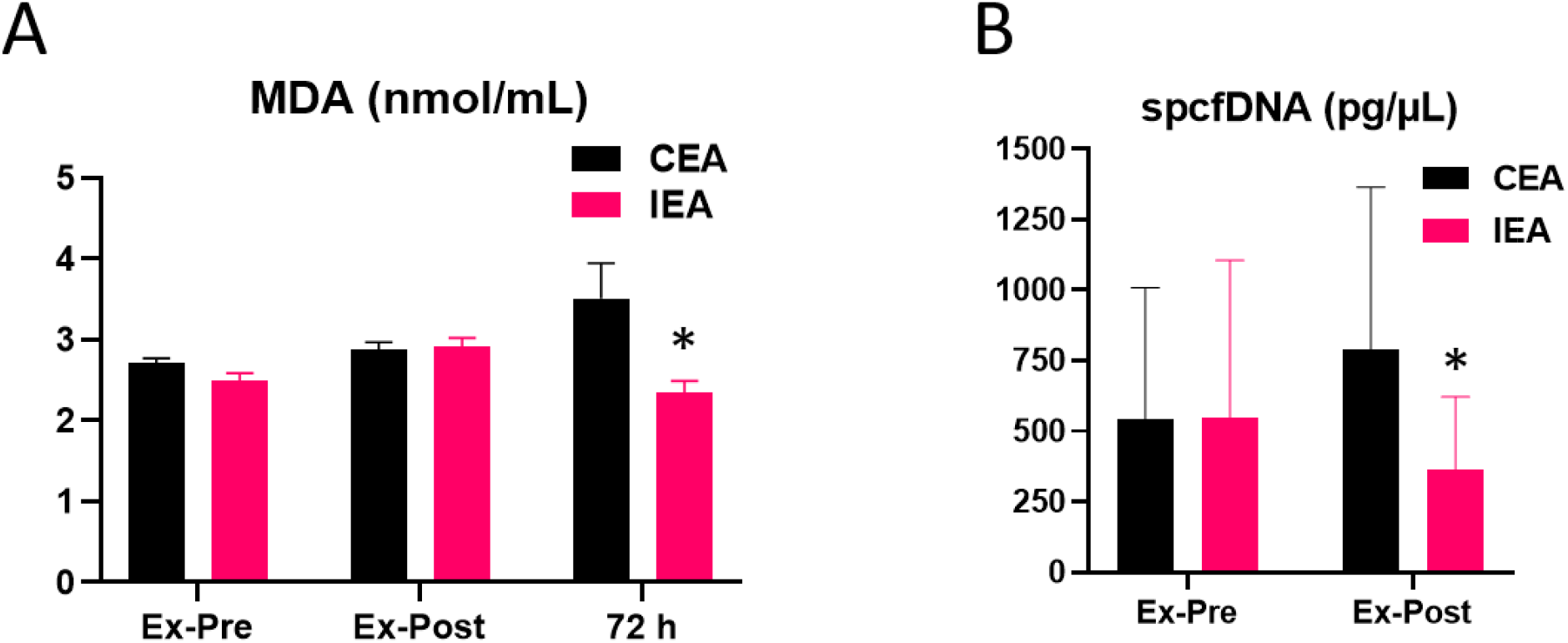
Oxidative stress marker and spcf DNA concentration. Changes in the serum concentrations of an oxidative stress marker and spcfDNA. (A) MDA concentration. (B) spcfDNA concentration. Asterisks indicate significant differences between groups (P < 0.05, Student’s t-test). MDA, malondialdehyde; spcfDNA, post-exercise-specific cell-free DNA; Ex-Pre, before exercise; Ex-Post, immediately after exercise; 72 h, 72 hours after exercise. n□=□10 in the CEA group, n = 9 in the IEA group.

## 4 Discussion

This study is the first to show that direct electroacupuncture muscle stimulation before and after exercise suppresses exercise-induced muscle soreness and damage. Additionally, the results revealed that electroacupuncture stimulation suppressed the increase in post-exercise levels of oxidative stress and spcfDNA concentration.

The IEA used in the present study suppressed DOMS after eccentric exercise. DOMS is known to occur 24–48 hours after eccentric exercise^24^. Additionally, several studies have reported the efficacy of the contralateral acupuncture technique^15, 25^. The present study found that IEA was more effective in suppressing DOMS than CEA. Direct stimulation may be involved in the inhibition of DOMS; however, this requires confirmation in further detailed analysis, including the use of a sham acupuncture group.

We found that muscle damage markers, such as creatine kinase and lactate dehydrogenase, appeared after the DOMS peak, as previously reported^10, 26, 27^, suggesting that the eccentric exercise was sufficient to induce damage. IEA suppressed DOMS after eccentric exercise but did not suppress the expression of markers of muscle damage. There are many uncertainties regarding the dynamics of muscle injury markers during muscle pain, and the concentrations of these markers may not align with the dynamics of muscle pain^28^. As electroacupuncture incorporates elements of electrical stimulation, it is conceivable that electroacupuncture may produce muscle protective effects similar to electrical stimulation^29^. However, electroacupuncture did not affect the concentrations of muscle injury markers. Electrical stimulation is considered to have the potential to influence neuropathic pain inhibition^30^; however, this concept requires further investigation.

Many studies have shown that the concentrations of lipid peroxidation products (MDA) are elevated in skeletal muscles and serum after transient exercise^31, 32^. An increased MDA concentration in the blood after eccentric exercise was previously reported to increase oxidative stress after 48–72 hours^27,32^. In the present study, electroacupuncture stimulation reduced the MDA concentration at 72 hours after exercise. A previous study showed that electrical pulse stimulation of C2C12 cells results in activation of the Nrf2 transcription factor, a master regulator of antioxidant enzymes^19^. Furthermore, sulforaphane supplementation for 2 weeks suppresses DOMS after eccentric exercise^27^. In the present study, it is possible that IEA stimulation may have increased the antioxidant capacity in the muscle and consequently suppressed the increase in the serum MDA concentration.

cfDNA fragments of various sizes leak into the blood as a result of cancer cell death, and analysis of these fragments has been reported to be a useful marker of cancer cells^33, 34^. Recently, cfDNA was analyzed as a marker of muscle damage shortly after exercising^35^. For example, the cfDNA concentration is increased immediately after weightlifting and high-intensity exercise (leg press)^11, 36^, while spcfDNA is reportedly a highly sensitive marker of extreme physical stress^12^. In the present study, IEA decreased the spcfDNA concentration immediately after exercise. Because electric current-based stimulation protects muscle cell membranes^29^, electroacupuncture stimulation may have inhibited cfDNA efflux from muscle tissue. Therefore, spcfDNA is presumed to flow out from the skeletal muscle, and this was prevented by electroacupuncture. Further research is required to confirm these findings.

### 4.1 Limitations

Our results suggest that electroacupuncture stimulation suppressed muscle soreness and damage after eccentric exercise. However, the present study did not include a sham electroacupuncture group. Furthermore, it remains unclear whether the suppression of muscle soreness was caused by electroacupuncture stimulation before (preventive effect) or after (therapeutic effect) the exercise.

## 5 Conclusion

IEA stimulation before and after eccentric exercise suppressed DOMS in young men and prevented an increase in the concentrations of muscle damage markers and spcfDNA compared with CEA. These results suggest the clinical effectiveness of electroacupuncture in preventing and treating sports-related injuries, thus enhancing the motivation to exercise in untrained individuals and those unfamiliar with exercise.

## Data Availability

All data produced in the present study are available upon reasonable request to the authors

## 6 Acknowledgments

We thank Benjamin Knight, MSc, Sydney Koke, MFA, and Kelly Zammit, BVSc, from Edanz (https://jp.edanz.com/ac) for editing a draft of this manuscript.

## 7 Funding

This study was supported by the JSPS KAKENHI (Grant Number: 22H03485) and Grant in Research Aid for Anna-Massage-Acupressure, Acupuncture and Moxibustion from the Public Interest Incorporated Foundation for Training and Examination in Anna-Massage-Acupressure, Acupuncture and Moxibustion.

## 8 Declaration of conflicting interests

The authors declare no competing interests.

## Notes

### Competing Interest Statement

The authors have declared no competing interest.

### Clinical Trial

UMIN000054171

### Author Declarations

Ethics Statement The study protocol was approved by the ethics committees of the University of Tsukuba, Japan (approval number: Tai019-75) and Teikyo Heisei University (approval number: 2020-100) and followed the ethics principles of the 7th revision (2013) of the Declaration of Helsinki. Participants provided written informed consent and authorization for the disclosure of protected health information before enrolling in the study.

## References

1. Basso JC, Oberlin DJ, Satyal MK, et al. Examining the Effect of Increased Aerobic Exercise in Moderately Fit Adults on Psychological State and Cognitive Function. Front Hum Neurosci 2022; 16: 833149. 20220712. DOI: 10.3389/fnhum.2022.833149.

2. Brito LC, Marin TC, Azevêdo L, et al. Chronobiology of Exercise: Evaluating the Best Time to Exercise for Greater Cardiovascular and Metabolic Benefits. Compr Physiol 2022; 12: 3621-3639. 20220629. DOI: 10.1002/cphy.c210036.

3. De Nys L, Anderson K, Ofosu EF, et al. The effects of physical activity on cortisol and sleep: A systematic review and meta-analysis. Psychoneuroendocrinology 2022; 143: 105843. 20220624. DOI: 10.1016/j.psyneuen.2022.105843.

4. Kagawa F, Yokoyama S, Takamura M, et al. Decreased physical activity with subjective pleasure is associated with avoidance behaviors. Sci Rep 2022; 12: 2832. 20220218. DOI: 10.1038/s41598-022-06563-3.

5. Liu D, Zhang Y, Wu L, et al. Effects of Exercise Intervention on Type 2 Diabetes Patients With Abdominal Obesity and Low Thigh Circumference (EXTEND): Study Protocol for a Randomized Controlled Trial. Front Endocrinol (Lausanne) 2022; 13: 937264. 20220712. DOI: 10.3389/fendo.2022.937264.

6. Cheung K, Hume P and Maxwell L. Delayed onset muscle soreness : treatment strategies and performance factors. Sports Med 2003; 33: 145–164. DOI: 10.2165/00007256-200333020-00005.

7. Proske U, Gregory JE, Morgan DL, et al. Force matching errors following eccentric exercise. Hum Mov Sci 2004; 23: 365–378. DOI: 10.1016/j.humov.2004.08.012.

8. Saxton JM, Clarkson PM, James R, et al. Neuromuscular dysfunction following eccentric exercise. Med Sci Sports Exerc 1995; 27: 1185–1193.

9. Weerakkody N, Percival P, Morgan DL, et al. Matching different levels of isometric torque in elbow flexor muscles after eccentric exercise. Exp Brain Res 2003; 149: 141-150. 20030125. DOI: 10.1007/s00221-002-1341-0.

10. Ra SG, Miyazaki T, Ishikura K, et al. Combined effect of branched-chain amino acids and taurine supplementation on delayed onset muscle soreness and muscle damage in high-intensity eccentric exercise. J Int Soc Sports Nutr 2013; 10: 51. 20131106. DOI: 10.1186/1550-2783-10-51.

11. Andreatta MV, Curty VM, Coutinho JVS, et al. Cell-Free DNA as an Earlier Predictor of Exercise-Induced Performance Decrement Related to Muscle Damage. Int J Sports Physiol Perform 2018; 13: 953-956. 20180728. DOI: 10.1123/ijspp.2017-0421.

12. Sugasawa T, Fujita SI, Kuji T, et al. Dynamics of Specific cfDNA Fragments in the Plasma of Full Marathon Participants. Genes (Basel) 2021; 12 20210430. DOI: 10.3390/genes12050676.

13. Itoh K, Ochi H and Kitakoji H. Effects of tender point acupuncture on delayed onset muscle soreness (DOMS)--a pragmatic trial. Chin Med 2008; 3: 14. 20081125. DOI: 10.1186/1749-8546-3-14.

14. Huang C, Wang Z, Xu X, et al. Does Acupuncture Benefit Delayed-Onset Muscle Soreness After Strenuous Exercise? A Systematic Review and Meta-Analysis. Front Physiol 2020; 11: 666. 20200717. DOI: 10.3389/fphys.2020.00666.

15. Guo Q, Di Z, Tian HF, et al. Contralateral Acupuncture for the Treatment of Phantom Limb Pain and Phantom Limb Sensation in Oncologic Lower Limb Amputee: A Case Report. Front Neurosci 2021; 15: 713548. 20211022. DOI: 10.3389/fnins.2021.713548.

16. Kim MK, Choi TY, Lee MS, et al. Contralateral acupuncture versus ipsilateral acupuncture in the rehabilitation of post-stroke hemiplegic patients: a systematic review. BMC Complement Altern Med 2010; 10: 41. 20100730. DOI: 10.1186/1472-6882-10-41.

17. Lu F. Clinical application of contralateral acupuncture technique. J Tradit Chin Med 1997; 17: 124–126.

18. Zhang S, Wang X, Yan CQ, et al. Different mechanisms of contralateral-or ipsilateral-acupuncture to modulate the brain activity in patients with unilateral chronic shoulder pain: a pilot fMRI study. J Pain Res 2018; 11: 505-514. 20180307. DOI: 10.2147/JPR.S152550.

19. Horie M, Warabi E, Komine S, et al. Cytoprotective Role of Nrf2 in Electrical Pulse Stimulated C2C12 Myotube. PLoS One 2015; 10: e0144835. 20151214. DOI: 10.1371/journal.pone.0144835.

20. Bassit RA, Pinheiro CH, Vitzel KF, et al. Effect of short-term creatine supplementation on markers of skeletal muscle damage after strenuous contractile activity. Eur J Appl Physiol 2010; 108: 945-955. 20091203. DOI: 10.1007/s00421-009-1305-1.

21. Clarkson PM, Nosaka K and Braun B. Muscle function after exercise-induced muscle damage and rapid adaptation. Med Sci Sports Exerc 1992; 24: 512–520.

22. Cooke MB, Rybalka E, Williams AD, et al. Creatine supplementation enhances muscle force recovery after eccentrically-induced muscle damage in healthy individuals. J Int Soc Sports Nutr 2009; 6: 13. 20090602. DOI: 10.1186/1550-2783-6-13.

23. Nosaka K and Newton M. Difference in the magnitude of muscle damage between maximal and submaximal eccentric loading. J Strength Cond Res 2002; 16: 202–208.

24. Ra SG, Choi Y, Akazawa N, et al. Effects of Taurine Supplementation on Vascular Endothelial Function at Rest and After Resistance Exercise. Adv Exp Med Biol 2019; 1155: 407–414. DOI: 10.1007/978-981-13-8023-5_38.

25. Zhang H, Sun J, Wang C, et al. Randomised controlled trial of contralateral manual acupuncture for the relief of chronic shoulder pain. Acupunct Med 2016; 34: 164-170. 20160121. DOI: 10.1136/acupmed-2015-010947.

26. Ispirlidis I, Fatouros IG, Jamurtas AZ, et al. Time-course of changes in inflammatory and performance responses following a soccer game. Clin J Sport Med 2008; 18: 423–431. DOI: 10.1097/JSM.0b013e3181818e0b.

27. Komine S, Miura I, Miyashita N, et al. Effect of a sulforaphane supplement on muscle soreness and damage induced by eccentric exercise in young adults: A pilot study. Physiol Rep 2021; 9: e15130. DOI: 10.14814/phy2.15130.

28. Baird MF, Graham SM, Baker JS, et al. Creatine-kinase-and exercise-related muscle damage implications for muscle performance and recovery. J Nutr Metab 2012; 2012: 960363. 20120111. DOI: 10.1155/2012/960363.

29. Vanderthommen M, Chamayou R, Demoulin C, et al. Protection against muscle damage induced by electrical stimulation: efficiency of a preconditioning programme. Clin Physiol Funct Imaging 2015; 35: 267-274. 20140429. DOI: 10.1111/cpf.12160.

30. Varela-Rodriguez S, Sanchez-Gonzalez JL, Sanchez-Sanchez JL, et al. Effects of Percutaneous Electrolysis on Endogenous Pain Modulation: A Randomized Controlled Trial Study Protocol. Brain Sci 2021; 11 20210617. DOI: 10.3390/brainsci11060801.

31. Oh-ishi S, Kizaki T, Ookawara T, et al. Endurance training improves the resistance of rat diaphragm to exercise-induced oxidative stress. Am J Respir Crit Care Med 1997; 156: 1579–1585. DOI: 10.1164/ajrccm.156.5.96-11035.

32. Paschalis V, Nikolaidis MG, Fatouros IG, et al. Uniform and prolonged changes in blood oxidative stress after muscle-damaging exercise. In Vivo 2007; 21: 877–883.

33. Hufnagl C, Leisch M, Weiss L, et al. Evaluation of circulating cell-free DNA as a molecular monitoring tool in patients with metastatic cancer. Oncol Lett 2020; 19: 1551-1558. 20191209. DOI: 10.3892/ol.2019.11192.

34. Pantel K and Alix-Panabieres C. Liquid biopsy and minimal residual disease - latest advances and implications for cure. Nat Rev Clin Oncol 2019; 16: 409–424. DOI: 10.1038/s41571-019-0187-3.

35. Vittori LN, Tarozzi A and Latessa PM. Circulating Cell-Free DNA in Physical Activities. Methods Mol Biol 2019; 1909: 183–197. DOI: 10.1007/978-1-4939-8973-7_14.

36. Atamaniuk J, Vidotto C, Kinzlbauer M, et al. Cell-free plasma DNA and purine nucleotide degradation markers following weightlifting exercise. European Journal of Applied Physiology 2010; 110: 695–701. DOI: 10.1007/s00421-010-1532-5.

